# Transient adverse events after REGN-CoV2 administration for mild COVID-19 patients and their potential predictive factors: a single center analysis

**DOI:** 10.1101/2021.11.29.21266623

**Authors:** Gen Kano, Kyoko Taniguchi, Yukiko Oue

## Abstract

**Background:** REGN-COV2, a monoclonal antibody cocktail drug against the SARS-COV-2 virus, has proven to be effective in preventing the development of severe COVID-19 and is increasingly being administered in outpatient and home settings. Adverse events such as fever and decreased oxygen saturation may occur after administration of REGN-COV2, and although these symptoms are generally mild and transient, predicting the occurrence of these adverse events is useful in developing a monitoring plan for patients.

**Methods:** We performed a retrospective analysis of 76 patients who received REGN-CoV2 between August and September 2021. We collected information on fever, decreased oxygen saturation requiring oxygen supplementation, and other adverse events from medical records. Patients were divided into two subgroups: those who presented with fever or oxygen desaturation and those who did not, and underlying medical conditions and laboratory data were compared between each group. The parameters that exhibited significant differences were further tested using Fisher’s exact test to evaluate whether appropriate thresholds could be set to distinguish the incidence group from the non-incidence group.

**Findings:** Of the 76 patients, 47 had fever of 38.5°C or higher within 24 hours after administration, and 27 of these patients had a body temperature of 37.5°C or lower before administration. Oxygen was required in 17 cases, 7 of which required oxygen more than 24 hours after administration of REGN-COV2, and additional treatment such as dexamethasone was given as the disease progressed to moderate. Among the parameters analyzed, lymphocyte count and IFNλ3 showed significant differences between the fever and non-fever groups. This was also the case in the comparison excluding patients who had fever before administration. There was also a significant difference in ferritin and CRP between the oxygen required and non-required groups. In addition to IFNλ3, ferritin, and CRP, there was a significant difference in LDH between the group that required additional treatment and the group that did not. When lymphocytes count <950/µL was used to predict fever, the sensitivity and specificity were 55% and 79%, respectively, with odds ratio 4.746 (95% CI: 1.666 to 14.12, p=0.004) in contingency table analysis. Similarly, when IFNλ3 >5.0 was used as the cutoff, sensitivity 72%, specificity 76%, odds ratio 8.220 (2.857 to 22.22; p<0.0001).

**Interpretations:** Transient fever and decreased oxygen saturation are common adverse events after REGN-CoV2 administration, and their occurrence correlated with the severity factor of COVID-19 itself. Evaluation of these items at the time of administration is useful not only for predicting the severity of COVID-19 but also for the development of adverse events in patients receiving REGN-CoV2.

## Introduction

Treatment with monoclonal antibodies has shown excellent efficacy in preventing the development of severe cases of COVID-19, and in countries with limited access to hospitalization, such as Japan, outpatient or home use is expected to reduce the demand for hospitalization. Although antibody therapy is a safe treatment, a certain number of patients have experienced adverse events such as transient high fever and decreased oxygen saturation after administration, which can lead to unscheduled medical visits, especially in patients who are not hospitalized, and is a burden for both patients and healthcare providers. If we can understand the trends of these adverse events and predict their occurrence, it will be easier to inform patients, reduce their anxiety, and curb unscheduled visits.

Therefore, we conducted a retrospective analysis of the incidence of adverse events in patients with mild COVID who received monoclonal antibody cocktail REGN-CoV2 at our institution and evaluated the factors that may predict the occurrence of adverse events.

## Methods

Patients with mild COVID-19 who were admitted to the Kyoto Yamashiro General Medical Center from August 2021 (immediately after the approval of REGN-CoV2 therapy in Japan) to September 2021 (until the so-called fifth wave due to the outbreak of the delta strain subsided in Japan) and treated with REGN-CoV2 (Ronapreve ® [casirivimab and imdevimab], Chugai Pharmaceutical Co., Ltd.) were included in the study. Patients who were mildly ill before treatment but later progressed to moderately ill or more were also included. Patient background (age, weight, BMI, smoking history, history of SARS-CoV-2 vaccination), symptom course (days from onset to administration, body temperature at admission), and hematobiochemical data (white blood cell, lymphocyte count, LDH, CRP, ferritin, TARC/CCL17, IFNλ3, HbA1c) were extracted from the medical records. The patients were classified into two groups according to the presence or absence of adverse events of interest (fever of 38.5°C or higher, decreased oxygen saturation requiring oxygen supplementation), and t-test was used to determine whether there were differences in the above parameters between the two groups. The parameters that showed significant differences were further evaluated their predictive value for the development of adverse events. For this purpose, patients are allocated to the 2×2 table with and without events and above and below to the threshold in a specific single parameter, and the significance of the association was analyzed by Fisher’s exact test. All statistical analyses were performed using Prism ® version 9.1 (Graphpad Software).

## Results

### Patient characteristics

Seventy-six patients were included in the study. The male to female ratio was 54:22, and the age ranged from 20 to 94 years (median 51.5 years). All patients had mild disease without oxygenation and within 7 days of onset (0-7 days, median 3days), both of which are mandatory for the use of REGN-COV2 at target period. At the time of admission, 53 patients had no fever or a low fever of 37.5°C or less.

REGN-COV2 is indicated for the treatment of patients with the following risk factors for developing severe disease in the U.S. Emergency Use Authorization (EUA): obesity >25 BMI, pregnancy, chronic renal disease, diabetes mellitus, immunodeficiency, cardiovascular disease or hypertension, and chronic lung disease including asthma. In addition, some patients met the inclusion criteria for the phase III study (COV-2067 study) (age 50 years or older) and items included as risk factors for the development of severe disease in Japan (malignancy, dyslipidemia). Most of the subjects had not been vaccinated, while 8 and 5 subjects had been vaccinated once and twice, respectively (**Table 1**).

**Table 1.**
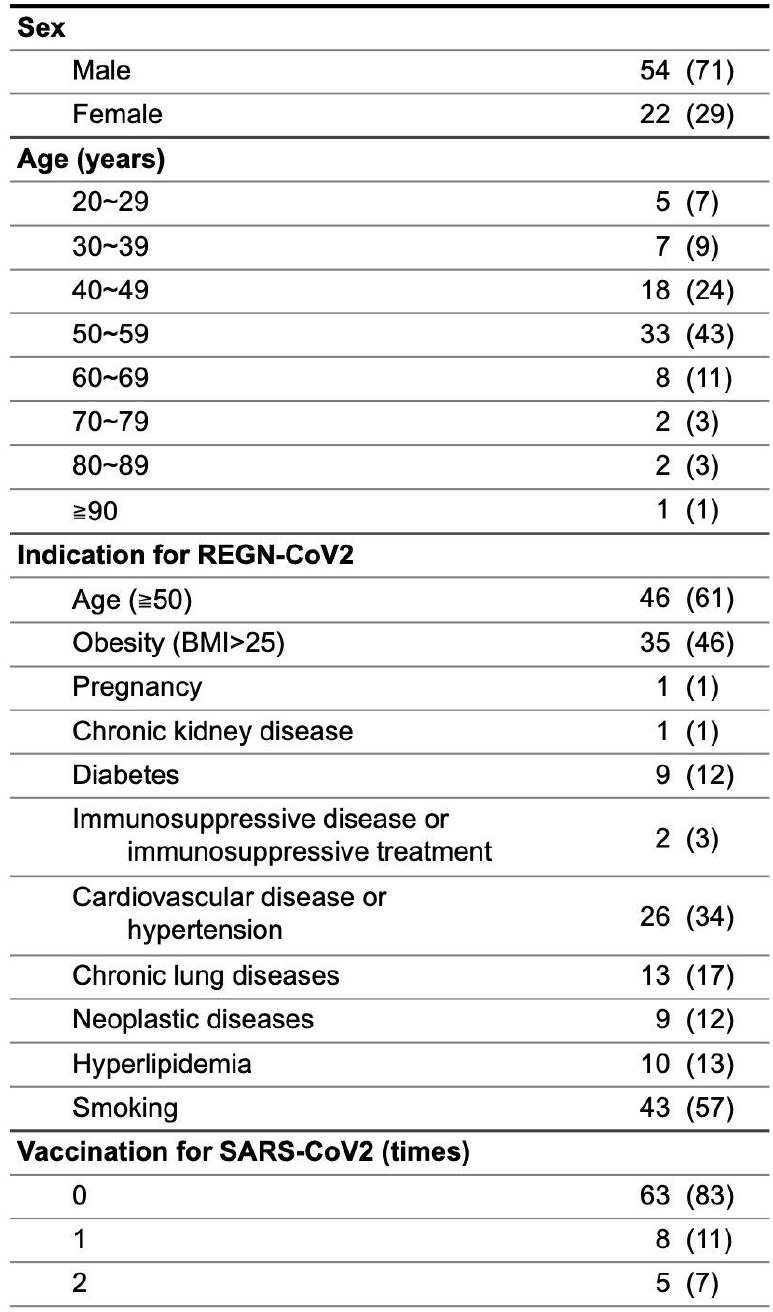
Characteritics of patients - no.(%)

| **Sex** |  |
| --- | --- |
| Male | 54 (71) |
| Female | 22 (29) |
| **Age (years)** |  |
| 20~29 | 5 (7) |
| 30~39 | 7 (9) |
| 40~49 | 18 (24) |
| 50~59 | 33 (43) |
| 60~69 | 8 (11) |
| 70~79 | 2 (3) |
| 80~89 | 2 (3) |
| ≥90 | 1 (1) |
| **Indication for REGN-CoV2** |  |
| Age (≥50) | 46 (61) |
| Obesity (BMI>25) | 35 (46) |
| Pregnancy | 1 (1) |
| Chronic kidney disease | 1 (1) |
| Diabetes | 9 (12) |
| Immunosuppressive disease or<br>immunosuppressive treatment | 2 (3) |
| Cardiovascular disease or<br>hypertension | 26 (34) |
| Chronic lung diseases | 13 (17) |
| Neoplastic diseases | 9 (12) |
| Hyperlipidemia | 10 (13) |
| Smoking | 43 (57) |
| **Vaccination for SARS-CoV2 (times)** |  |
| 0 | 63 (83) |
| 1 | 8 (11) |
| 2 | 5 (7) |

### Adverse events

Of all 76 patients, 47 experienced a fever of 38.5°C or higher within 24 hours after administration of REGN-CoV2 (**Figure 1A, B & C**). 17 required oxygen administration due to decreased oxygen saturation, but 10 of these patients were able to finish oxygen within 48 hours. The remaining 7 patients progressed to moderate disease and required additional treatment with dexamethasone or other agents. When limited to 53 patients who were afebrile (37.5°C or less) at the start of REGN-COV2 administration, 27 patients (51%) developed fever after administration, 14 patients (26%) required oxygen administration, and 6 patients (12%) progressed to moderate disease and required additional treatment. (**Table 2**). None of the patients who transitioned to moderate disease became severely ill to require HFNC or ventilator management, and all were discharged within 14 days. Other minor adverse events included headache: 23 patients (30%), nausea: 10 patients (14%), and diarrhea: 7 patients (9%) within 24 hours.

**Table 2.**
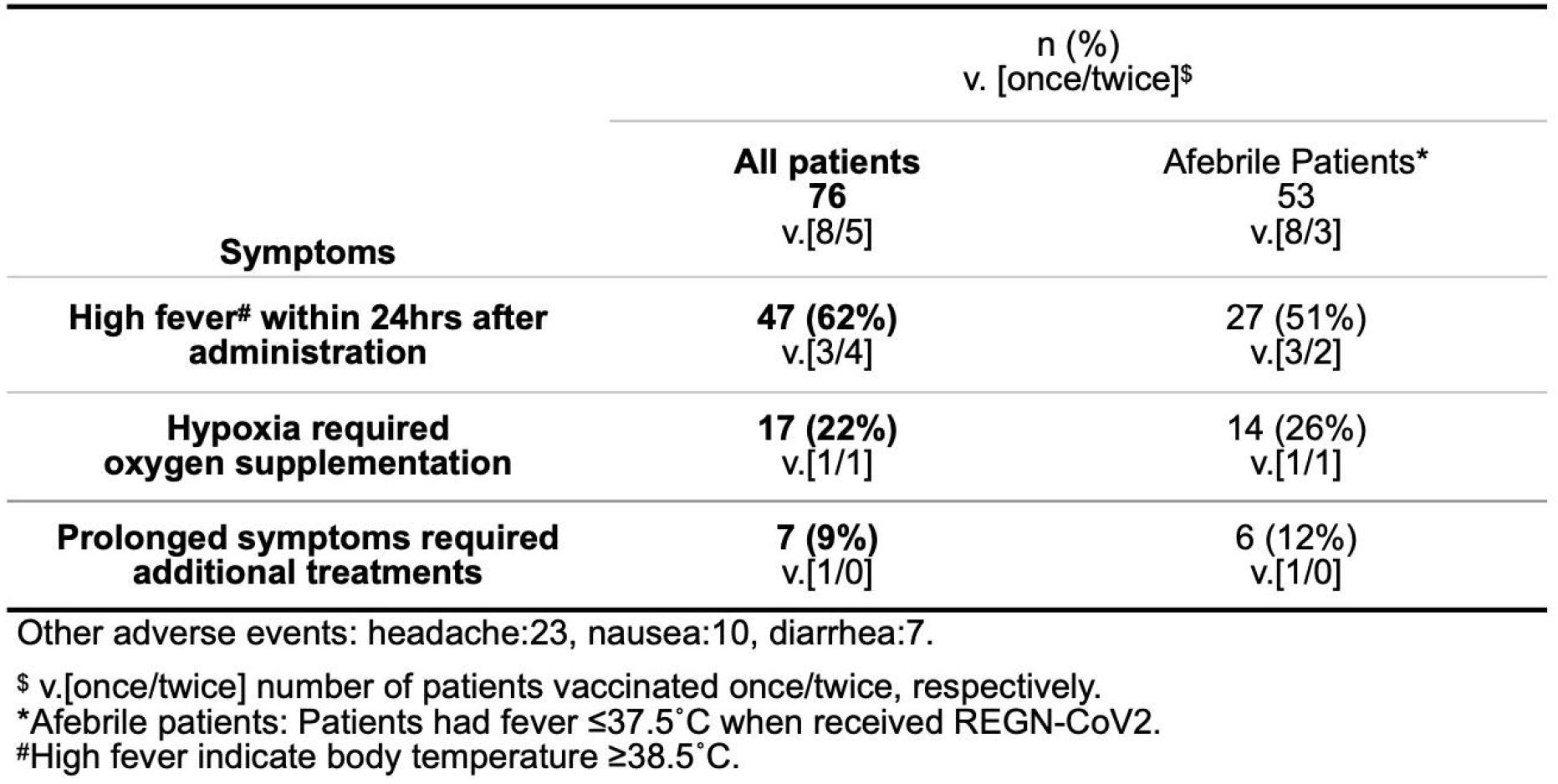
Adverse events after REGN-CoV2 administration.

| **Symptoms** | **n (%)**<br>**v. [once/twice]<sup>\$</sup>** | |
| --- | --- | --- |
|  | **All patients**<br>**76**<br>**v.[8/5]** | **Afebrile Patients\***<br>**53**<br>**v.[8/3]** |
| **High fever<sup>#</sup> within 24hrs after administration** | **47 (62%)**<br>**v.[3/4]** | **27 (51%)**<br>**v.[3/2]** |
| **Hypoxia required oxygen supplementation** | **17 (22%)**<br>**v.[1/1]** | **14 (26%)**<br>**v.[1/1]** |
| **Prolonged symptoms required additional treatments** | **7 (9%)**<br>**v.[1/0]** | **6 (12%)**<br>**v.[1/0]** |

Other adverse events: headache:23, nausea:10, diarrhea:7.
<sup>\$</sup> v.[once/twice] number of patients vaccinated once/twice, respectively.
\*Afebrile patients: Patients had fever $\leq 37.5^{\circ}\text{C}$ when received REGN-CoV2.
<sup>#</sup>High fever indicate body temperature $\geq 38.5^{\circ}\text{C}$ .

**Figure 1.**
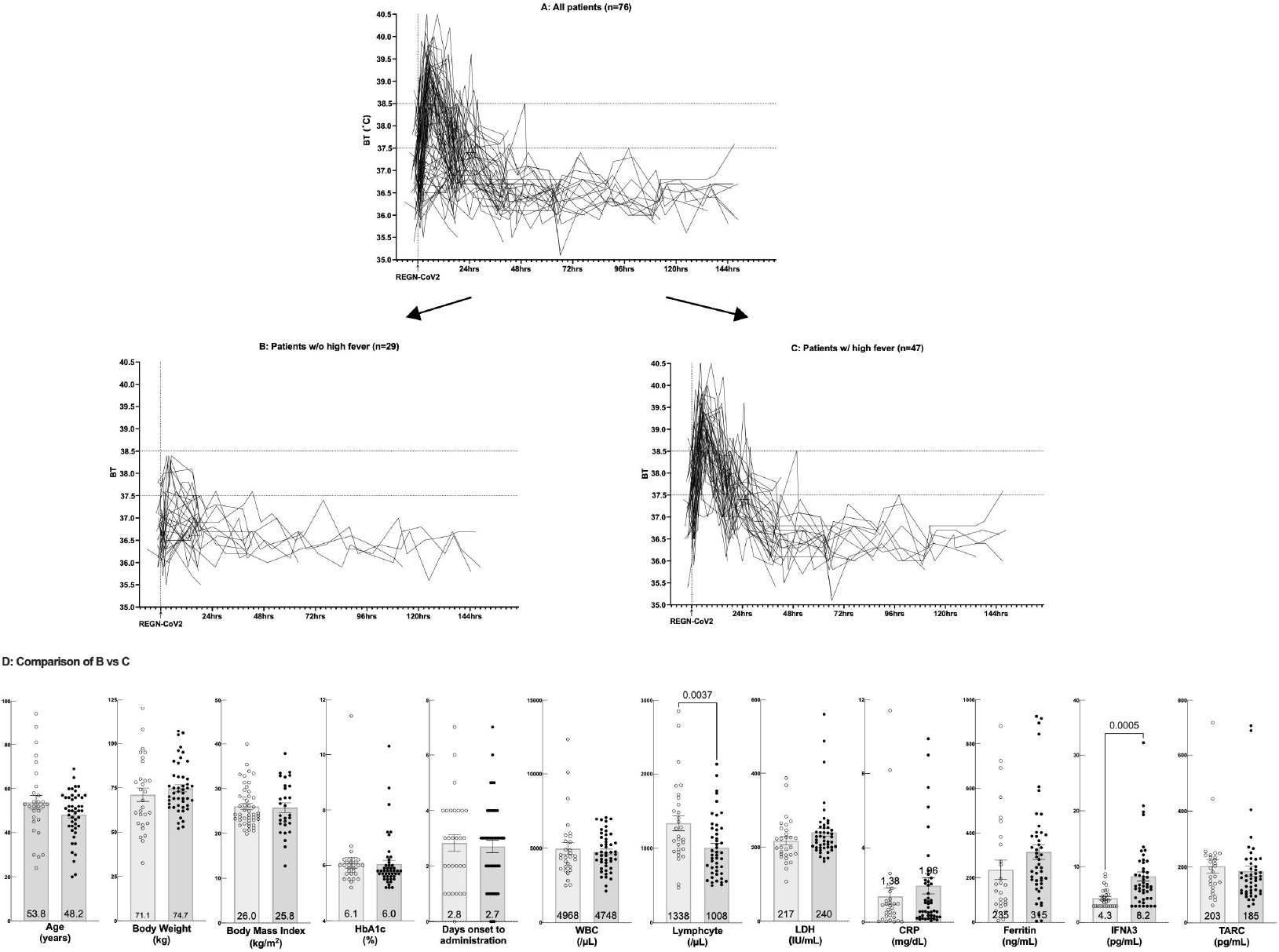
Temperature chart and comparison of parameters with and without the high fever. A. Temperature chart of all patients (n=76) based on the starat time of REGN-CoV2 administration (arrow). B. Temperature chart only showing patients who did not experience high fever (≥38.5°C) after administration of REGN-CoV2 (n=29). C. Temperature chart only showing patients who experienced high fever (≥38.5°C) after administration of REGN-CoV2 (n=47). D. Comparison of parameters in patients without high fever (open circles) and patients with high fever (closed circles). Each bar represent mean with error bar showing standard deviation (mean values are labeled on the bar). P-values are only displayed when the t-test yields a value of <0.05.

### Parameters Comparison the adverse event vs the non-event group

We examined whether there were differences in various parameters of adverse events, such as fever and decreased oxygen saturation, between the incidence and non-incidence groups. The results showed that the lymphocyte count was significantly lower in the group with fever than in the group without fever (1008 ±405.8 µL vs 1338 ±550.3 µL, respectively; p=0.0037). On the other hand, IFNλ3 was significantly higer in fever group compated to no-fever froup (8.234 ±5.668 pg/mL vs 4.321 ±1.688 pg/mL, respectively; p=0.0005). Other parameters, namely age, body weight, BMI, HbA1c, days from onset to administration, WBC, CRP, LDH, ferritin and TARC, showed no significant diffrence between both groups (**Figure 1D**). Since it is obvious to some extent that patients who had fever before administration would have high fever, we excluded these patients from the comparison, but still found significant differences in lymphocyte count and IFNλ3 (**Supplemental figure 1**). In addition, to prevent bias due to extreme values in patients with moderate disease, we excluded these patients from the comparison and found that IFNλ3 was still significantly higher in the fever group (**Supplemental figure 2**). Patients who required oxygen had significantly lower lymphocyte counts (882 ±291.6 µL vs 1206 ±512.9 µL; p=0.0152) and higher levels of CRP (4.591 ±4.527 mg/dL vs 0.916 ±1.114 mg/dL; p<0.0001), ferritin (502.7 ±302.8 ng/mL vs 221.8 ±160.1 ng/mL; p<0.0001), and IFNλ3 (10.18 ±8.387 pg/mL vs 5.751 ±2.779 pg/mL; p=0.0008) than those who did not (**Figure 2A**). When comparing patients who developed moderate disease and required add-on treatments, CRP (6.329 ±5.118 mg/dL vs 1.272±1.950 mg/dL; p<0.0001), ferritin (586.1 ±318.0 ng/mL vs 254.0 ±198.8 ng/mL; p=0.0002), LDH (332.7 ±155.4 IU/L vs 221.0 ±47.30 IU/L; p<0.0001) and IFNλ3 (15.56 ±8.987 pg/mL vs 5.846 ±3.317 pg/mL; p<0.0001) were significantly higher than patients did not require add-on (**Figure 2B**).

**Figure 2.**
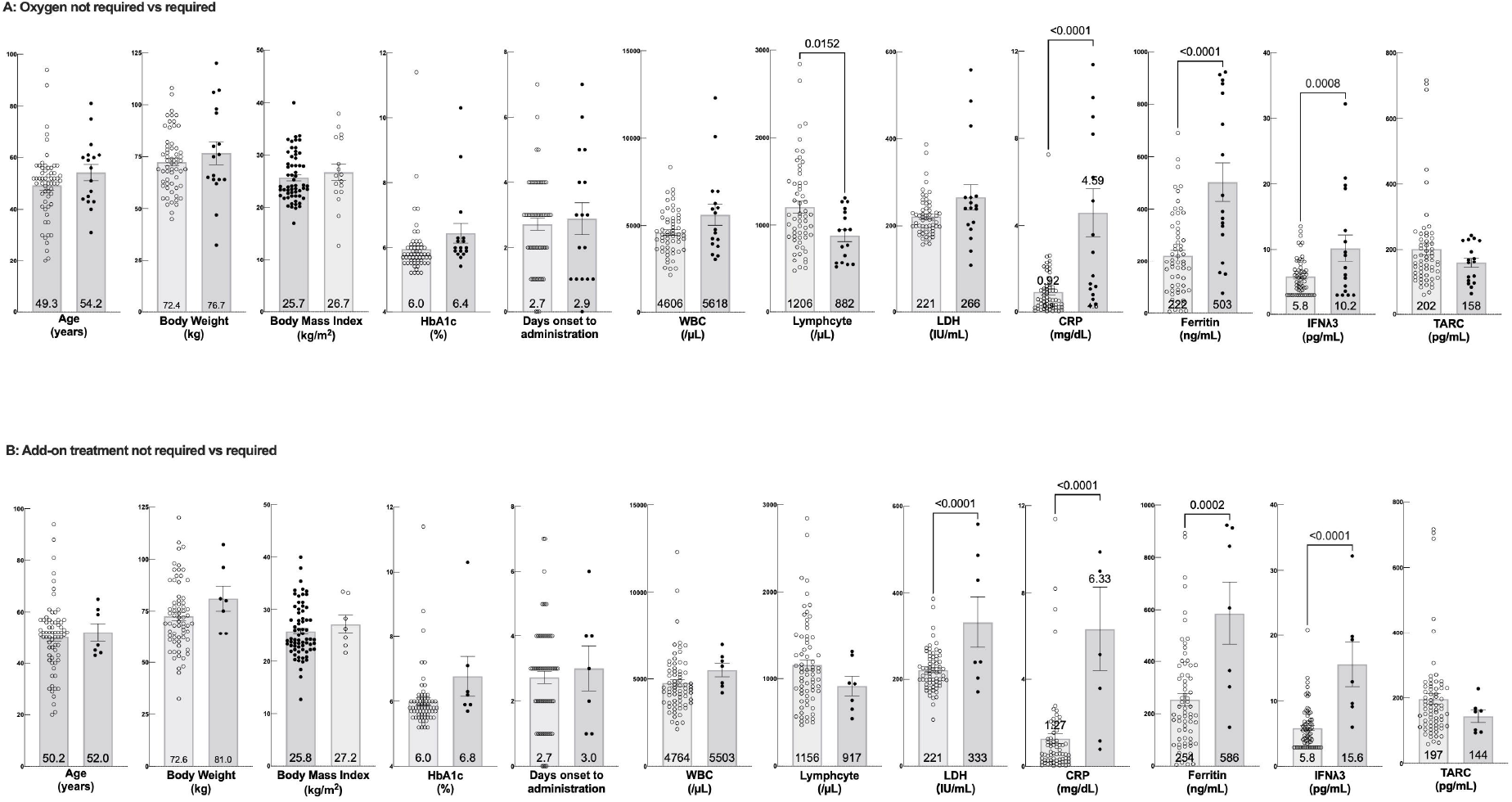
Parameter comparisons for oxygen requirement and add-on threatment requirement. A. Comparison of patients who did not required oxygen supplementation (n=59; open circles) vs patients who did not (n=17, closed circles) B. Comparison of patients who did not required add-on treatment (n=69; open circles) vs patients who did not (n=7, closed circles) Each bar represent mean with error bar showing standard deviation (mean values are labeled on the bar). P-values are only displayed when the t-test yields a value of <0.05.

### Validation of predicting potential for each parameter

Next, we conducted simulations to see if we could predict the occurrence of adverse events by setting appropriate thresholds for these parameters that showed significant differences. Fisher’s exact test demonstrated that a threshold of less than 950 lymphocytes resulted in a sensitivity of 55% and specificity of 79%, with odds ratio of 4.746 for fever (p=0.004). Similarly, cut-off calue of IFNλ3>5.0 yielded sensitivity 72% and specificity 76%, with odds ratio 8.220 (p<0.0001).

For oxygen requirement, each of lymphocyte count, CRP, ferritin and IFNλ3 was found to have proper threshold (see **Figure 3**) to predict the occurrence of the event. CRP, LDH, ferritin, and IFNλ3 were also demonstrated to be useful in predicting the need for additional treatment by certain cut-off values (**Figure 3**).

**Figure 3.**
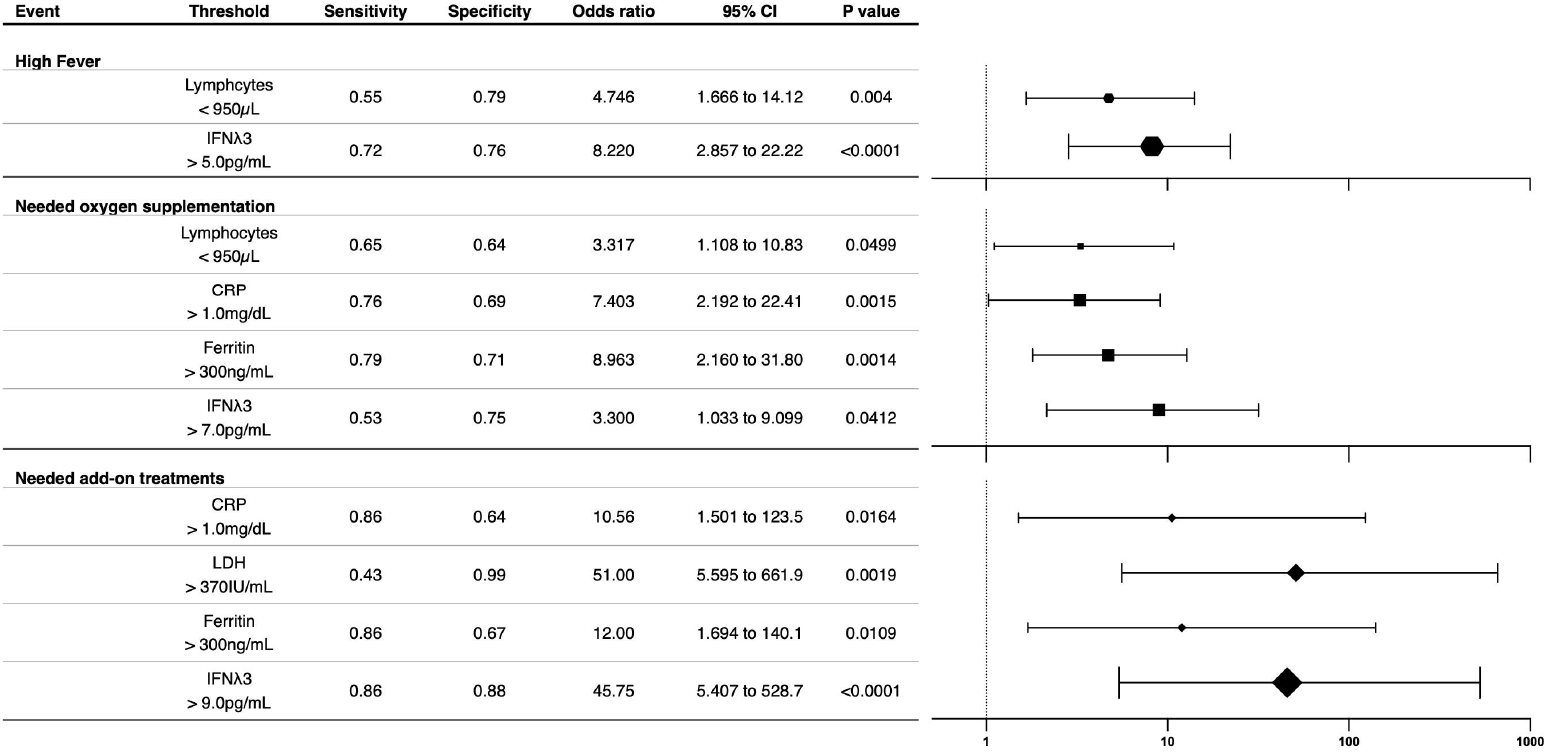
Examining the diagnostic value of each parameter. Based on the 2 × 2 tables generated by setting shown thresholds for each parameter, their diagnostic value was examined by Fisher exact test. Odds ratios and 95% confidence intervals are shown as Forrest plots, with the size of the dot reflecting the high confidence level inferred from the P value.

## Discussion

In our cohort, fever after REGN-CoV2 administration was observed in a high percentage of patients (56%; 47/76). Even among patients who did not have fever before treatment, a relatively high percentage (40%; 21/53) developed high fever immediately after treatment and resolved within 48 hours, indicating a strong temporal correlation with treatment. The risk of fever was higher in patients with low lymphocyte counts and high IFNλ3 levels. In addition to this, high ferritin and CRP were associated with the need for oxygen. In addition, if LDH was also high, patients were more likely to require additional treatment (transition to moderate disease). IFNλ3 is an antiviral cytokine, and high levels of IFNλ3 have been reported to be a predictor of early stage of severe COVID-19 [Ref1]. In this study, we showed that IFNλ3 can be a very sensitive indicator in predicting transient adverse events after REGN administration. It is not yet widely used as a point-of-care test with immediate results, however, which makes it difficult to apply this test in outpatient or home treatment settings. In such situations, it is better to make a comprehensive judgment based on lymphocyte count and other factors.

REGN-CoV2 was approved for emergency use by the FDA on November 21, 2020 as a drug for the prevention of severe disease in patients with COVID-19, and has since been used in many countries including the UK, EU, and Japan [Ref2]. Patients receiving REGN-Cov2 have been shown to have a 71.3% reduction in hospitalization rates and a 70.4% reduction in mortality compared to patients receiving placebo [Ref3]. REGN-CoV2 is basically a well-tolerated drug, as the Global Phase 1/2/3 Randomized Placebo-controlled Study (COV-2067, overseas data) reported 1.1% serious adverse events (including severe hypersensitivity reactions, infusion reactions) [Ref4]. The adverse events observed in this study, such as fever, were not accompanied by other symptoms characteristic of infusion reactions, such as hypotension and rash, and it is highly debatable whether the transient fever or mild hypoxia found in our study should be included in infusion reactions. The fact that the adverse events observed in this study were associated with known COVID-19 severity factors such as lymphocyte count and IFNλ3, rather than underlying conditions such as age and body mass index, suggests that these adverse events are attributable to COVID-19 disease activity itself. REGN-CoV2 has already been reported to exacerbate symptoms in severely affected patients requiring high volume oxygen or mechanical ventilation [Ref5]. Although there is no detailed mention of the mechanism of this worsening of symptoms, it is assumed that REGN-COV2 may be involved in the abnormal immune activation in the process of developing severe COVID-19. The present results may indicate that REGN-CoV2 may lead to the manifestation of symptoms, even if only temporarily, even when administered to patients with mild disease. If so, the degree of deterioration may depend on the amount of virus present at the timing of REGN-CoV2 administration. Prophylactic administration of REGN-CoV2 to uninfected patients is expected to be expanded in the future, and if it is observed that uninfected patients do not develop fever after administration of REGN-CoV2, this may be a reason to suspect a relation to viral load, and it would be interesting to focus on this in order to consider not only adverse events of REGN-CoV2 but also the mechanism of the development of severe disease of COVID-19.

## Conclusion

With the antibody cocktail REGN-CoV2, even patients with mild disease are prone to transient fever and decreased oxygen saturation, and the risk factors for these events are similar to those for severe disease with COVID-19. More detailed investigation of these adverse events will lead to better patient management, and investigation of the underlying mechanisms may also provide insight into the mechanisms of the development of severe disease in COVID-19.

## Data Availability

All data produced in the present study are available upon reasonable request to the authors.

