## Supplementary figures and images for "Transient adverse events after REGN-CoV2 administration for mild COVID-19 patients and their potential predictive factors: a single center analysis"

### Supplemental figure 1

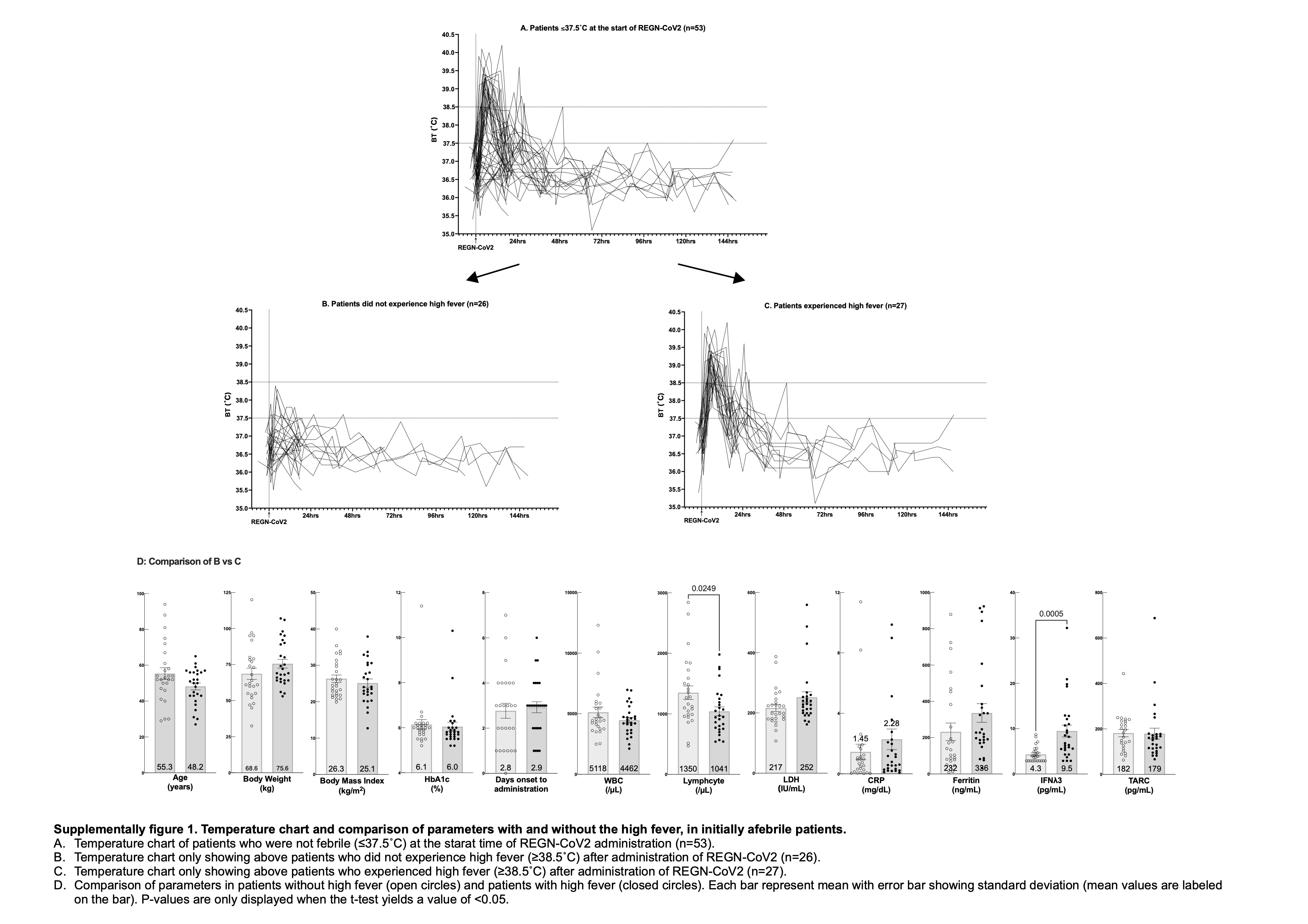

### Supplemental figure 2

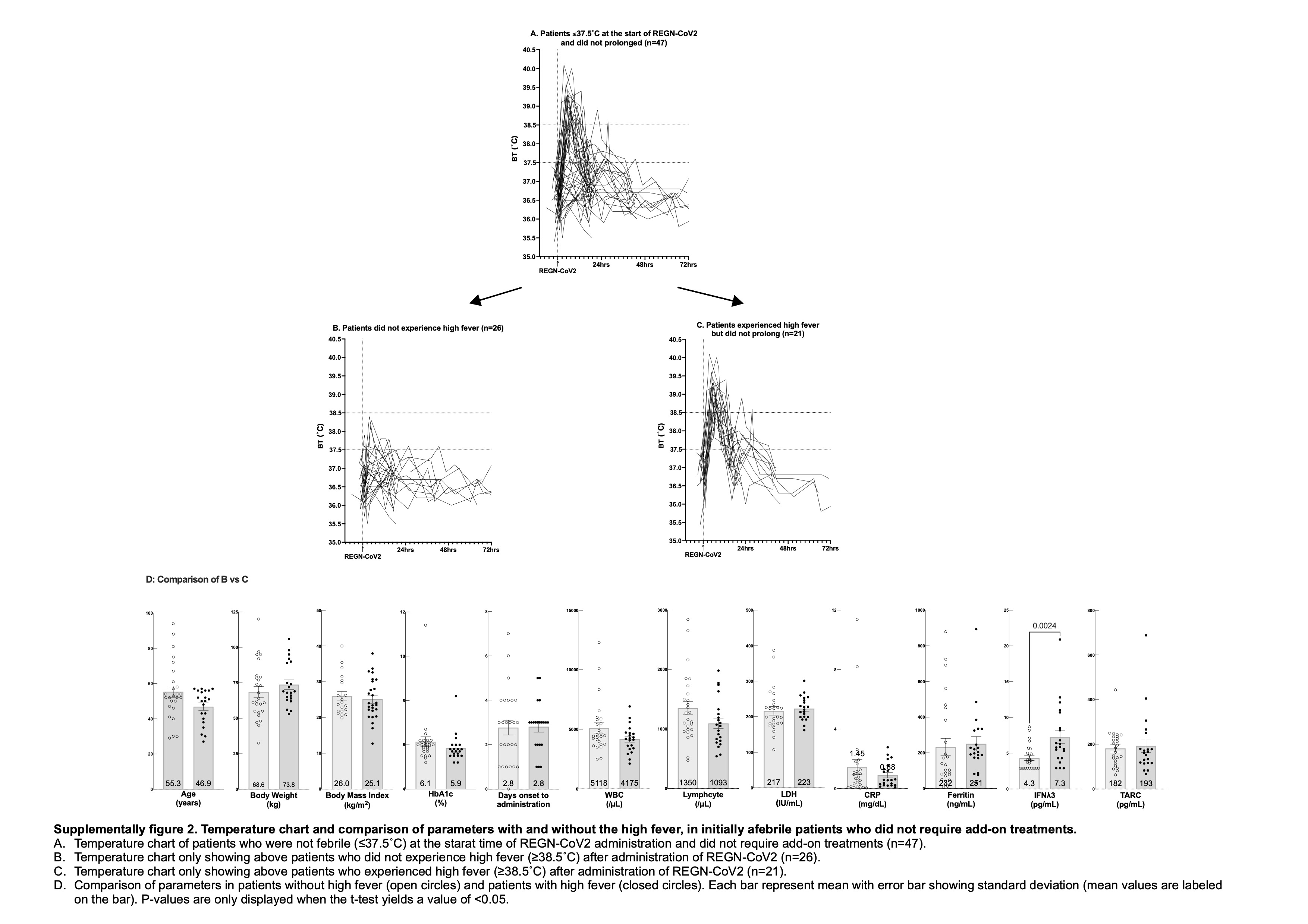
